# Development of HPV 16/18 E6 oncoprotein paper-based nanokit for enhanced detection of HPV 16/18 E6 oncoprotein in cervical cancer screening

**DOI:** 10.1101/2020.04.29.20084459

**Authors:** Lucy Muthoni Mwai, Mutinda C. Kyama, Caroline W. Ngugi, Edwin Walong

## Abstract

Cervical cancer caused mainly by high risk human papillomavirus (HPV) 16 and 18 strains is the second most prevalent cancer of women in Kenya. It is often diagnosed late when treatment is difficult due to very low percentage of women attending screening thus, mortalities remain high. The most available tests in low-and-middle-income countries (LMICs) have relatively low specificity, low sensitivity, require a laboratory setting and huge technical and financial support not readily available. HPV 16/18 E6 oncoprotein has been identified as a potential biomarker in a more specific early diagnosis of cervical cancer. This retrospective cross-sectional study developed a paper-based nanokit with enhanced detection of HPV 16/18 E6 oncoprotein for cervical cancer screening. The HRP labelled antibodies HPV 16 E6/18 E6-HRP (CP15) passively conjugated to citrate stabilized 20nm gold nanoparticles were evaluated for immune sensing mechanism using a recombinant viral HPV E6 protein. The diagnostic accuracy was evaluated using 50 tissue lysates from formalin fixed paraffin embedded cervical biopsy, including control (n=10), Mild Dysplasia (n=10), Cervical intraepithelial neoplasia 3 (CIN3) (n=10), Cervical intraepithelial neoplasia 4 (CIN4) (n=10) and invasive carcinoma (n=10). The molecular technique used was dot blot molecular assay. A positive result was generated by catalytic oxidation of peroxidase enzyme on 3,3’,5,5’-Tetramethylbenzidine (TMB) substrate. The gold nanoparticles were used to enhance the signal produced by peroxidase activity of horseradish peroxidase (HRP) enzyme giving a more sensitive assay as compared to use of non-conjugated antibody. This study provides a significantly high and reliable diagnostic accuracy for precancerous and cancerous lesions with a sensitivity of 90%, a specificity of 90%, a likelihood ratio for positive and negative tests as 9:1 and 1:9 respectively, a Positive Predictive Value of 97.3% and a Negative Predictive Value of 69.2%. This study avails a sensitive, rapid test using paper-based nanotechnology which can be utilised in community-based screening outreaches particularly in low- and middle-income countries.

## Introduction

According to global cancer statistics GLOBOCAN, carcinoma of cervix is ranked as the fourth most common malignancy among women worldwide with an estimation of 570,000 cases and 311,000 deaths in 2018 (1). It is the second most common female malignancy in Low-and-Middle Income Countries (LMICs). In Kenya, the prevalence is 25 cases per 100,000 women (2,3). Approximately 75% cases of cervical cancer are caused by persistent infections of the cervical mucosal epithelium with carcinogenic types of human papillomaviruses (HPVs) mainly 16 and 18 (4).

The gold standard test for cervical cancer diagnosis is histopathology that uses tissue biopsies (5). Early screening is a critical intervention point especially in Kenya, where most cancers are often diagnosed at advanced stage (2). WHO recommends screening at least once annually for all women between 30–65 years. The most recommended tests include: visual inspection VIA/VILI test, Papanicolaou (Pap) cytology and HPV testing (6).

The visual inspection with acetic acid (VIA) and visual inspection using Lugol’s iodine (VILI) are visual cervical cancer screening approaches. VIA examines the cervix after application of 3-5% acetic acid; if the cervical epithelium has abnormal cellular proteins, the acetic acid will coagulate them hence is visualized as an opaque and white area called acetowhite (7). From a study in China, the sensitivity of VIA test was found as 54.6% with a specificity of 89.9% for Cervical intraepithelial neoplasia 3+ (CIN3+) (8). The VILI test examines the cervix after application of Lugol’s iodine which reacts with glycogen present in normal mature squamous epithelium to form a brown-black coloration. In the presence of precancerous lesions and cancer which have little or no glycogen a yellow colour is observed (7).

Pap cytology involves staining of exfoliating cells from the transformation zone of the cervix with Papanicolaou stain, followed by microscopic detection of cancerous or precancerous lesions (9). It has a poor clinical sensitivity of 51% (10). It is subjective due to poor intraobserver and interobserver reproducibility in discriminating high-grade intraepithelial lesions, HSIL (CIN2/3) from low-grade intraepithelial lesions, LSIL (CIN1) due to dependence on cytologist’s microscopic interpretation of distribution and extent of abnormal cells (11). Additionally, successful Pap requires repeats causing substantial cost burdens to patients and the general economy (12). Finally, pap diagnosis of minor cytological lesions namely, equivocal atypical squamous cells of undetermined significance (ASCUS) and mildly abnormal LSIL poses a huge challenge to detect possible underlying CIN3 (13). Due to these challenges, pap cytology alone may not be effective in proper diagnosis and triage.

HPV tests identify the presence of HPV. In a study on Chinese women, HPV DNA test yielded a relatively high sensitivity for CIN 2+ at 70% and CIN3+ at 80% (14,15). HPV DNA has low specificity due to its inability to discriminate transient infections from persistent infections (8). New tests to determine the expression levels of HPV transcripts (E5, E6 and E7 mRNA) and proteins (E5, E6 and E7) connected to oncogenic HPV are being utilized. Their concentration is lower during transient infections hence better diagnostic value (16). A study on E6 protein determined the sensitivity and specificity of HPV16/18/45 E6 protein as 42.8% and 94.3% for CIN2+ respectively and 54.2% and 93.8% for CIN3+ respectively (14). Though relatively lower sensitivity, the specificity is commendable to discriminate transient infections.

HPV 16/18 E6 oncoprotein has been evaluated as a useful biomarker with prognostic abilities as it can detect pre-cancer and cancerous states of cervical cancer progression (17–19). A positive E6 assay, indicates high correlation to the cervical cancerous phenotype, not the potential for cervical cancer, thus high specificity in triaging patients during screening (16,20,21). E6 levels of expression associate directly with severity of CIN lesions and risk of progression to invasive carcinoma (22). Although, the current interventions utilizing the E6 oncoprotein from high risk HPV strains have high specificity, the sensitivity is relatively low. Arbor Vita Corporation’s OncoE6 test showed 99.0% specificity and 69.6% sensitivity for HSIL (CIN3+) precancerous lesions (19). HPV16/18/45 E6 test produced sensitivity and specificity of CIN2+ as 42.8% and 94.3% respectively and for CIN3+ as 54.2% and 93.8% respectively (4,14). More advanced methods of detecting E6 oncoprotein are therefore required to produce a sensitivity between 76.3 % -97.2% which is indicative of diagnostic test quality (23).

Gold nanoparticle (AuNP) bioconjugates are increasingly being utilised in biomedicine due to their low toxicity on biological tissues and unique electronic and chemical properties. In a recent study, the bioconjugation of AuNPs with HPV 16 E6/18 E6-HRP (CP15) antibody against HPV 16/18 E6 oncoprotein that is overexpressed in cervical carcinoma progression was developed through physical adsorption (25).

This study therefore, developed a paper-based nanokit through evaluation of the immune sensing mechanism and diagnostic accuracy of the HPV 16 E6/18 E6-HRP (CP15)-AuNPs bioconjugate in enhanced detection of E6 oncoprotein in cervical cancer screening.

## Materials and methods

### Study design

The study was a proof of concept retrospective cross-sectional study carried out at PAUSTI Molecular Biology and Biotechnology Laboratory and Histopathology Laboratory of the University of Nairobi, Kenya.

### Study species

The study utilised deidentified archived cervical formalin-fixed paraffin embedded (FFPE) biopsy tissue samples of confirmed pathology (n = 50) from the University of Nairobi, Histology Department FFPE repository. The study selected 51 formalin fixed paraffin embedded biopsy samples, including control/negative tissues (n= 10), Mild Dysplasia tissues (n=10), CIN3 tissues (n=10), CIN4 tissues (n=10) and invasive carcinoma tissues (n=11) and from a population of 100 deidentified cervical tissue based on published tables with a specified criteria where precision level is ±10%, confidence level 95% and P=0.5 (24). The samples were selected using purposive non-probability method of sampling. All samples selected were collected from women aged 18 years and above which had confirmed histopathology as negative or with precancerous lesions or cervical cancer lesions. Since archived samples were utilised, the age of FFPE used were collected and analysed less than 10 years prior to this investigation (between 2012–2014), samples with bigger tissue sizes were preferred where possible. Cervical tissues samples from women with other type of malignancies, or on treatment for cervical cancer, or those who had undergone surgery or those whose histopathology diagnosis varied between the resident and consultant pathologist or those whose amount of tissue was considerably small were excluded.

### Ethical statement

The study protocol was approved by the institutional Jomo Kenyatta University of Agriculture and Technology (JKUAT) ethical and research committee.

### Evaluation of immune sensing mechanism of the HPV 16 E6/18 E6-HRP (CP15) -AuNPS bioconjugate

#### Dot Blot Molecular Technique

Paper strips of 0.22μm pall nitrocellulose membrane (NC) (Thermofisher) and 125mm No. 1 Macherey-Nagel filter papers (MN) (Macherey-Nagel GmbH & Co. KG Germany Cat# MN 615.Ɵ) were cut using a pair of sterilized scissors and handled with surgical gloves and tweezers to avoid any surface alteration. A grid was drawn on the NC and MN strips using a pencil to indicate the region of the blot. A pelican HB pencil was used for marking and numbering the NC and MN strips. The recombinant viral HPV protein (Lot # 25158317A, Cat. # AP-120, 0.5mg/ml bio-techne an R&D Systems Company) was diluted into the concentrations indicated in Table 1.

**Table 1.** Showing differing dilutions of recombinant viral HPV E6 purified protein using a protein diluent comprising of 1X PBS, 400mM NaCl and 0.001M DTT.

| **Tube No.** | **Concentration of E6 protein (mg/ml)** | **Amount of E6 Protein stock (µl)** | **Amount of protein Diluent (µl)** |
| --- | --- | --- | --- |
| 1. | 0.020 | 4 | 996 |
| 2. | 0.006 | 12 | 988 |
| 3. | 0.010 | 20 | 980 |
| 4. | 0.014 | 28 | 972 |
| 5. | 0.018 | 36 | 964 |
| 6. | 0.022 | 44 | 966 |
| 7. | 0.026 | 52 | 948 |
| 8. | 0.030 | 60 | 940 |
| 9. | 0.034 | 68 | 932 |
| 10. | 0.038 | 76 | 924 |
| 11. | 0.042 | 80 | 920 |

To ensure optimization, small spots of 2 μl of the different concentrations of the recombinant viral HPV E6 protein sample were spotted onto the NC and MN paper strips at the center of the grid slowly ensuring about 2–4 mm diameter using narrow-mouth FinTip pipette tips (Thomas Scientific). The paper strips were air dried and non-specific sites blocked by soaking in 5% BSA-V in 1X TBS-T (Cat No. T1085 Beijing Solarbio Science & Technology Co. Ltd) and in a 10 cm Petri dish for 1 hour at room temperature. The membranes were incubated with HPV 16 E6/18 E6-HRP (CP15)-AuNPs bioconjugate (25) to allow formation of an immune complex at a concentration of 4 μg/ml for 30 min at room temperature. The paper strips were washed three times each about 5 minutes with TBS-T and incubated with TMB reagent (Thermofisher). A timer was started on addition of TMB reagent to determine the time of color development in positive reactions. A colour change was observed in 3 minutes.

#### Direct Deposit on nitrocellulose membrane and filter paper strips

About 5 μl of the different concentrations of the recombinant viral HPV E6 protein samples a range of 0mg/ml to 0.012mg/ml, were placed in different eppendorf tubes. 5μl HPV 16 E6/18 E6- HRP (CP15)-AuNPS bioconjugate was added to allow immune complex formation for 5 minutes. 2 μl of the immune complex was spotted onto the paper strips immobilized with TMB substrate and air dried. A colour change was generated by catalytic oxidation of peroxidase enzyme on 3,3’,5,5’-Tetramethylbenzidine (TMB) substrate in 3 minutes.

### Determination of detection limit using the dot blot molecular technique

The dot blot molecular technique was carried out using serially diluted recombinant viral HPV E6 protein into a protein diluent comprising 1X PBS (Beijing Solarbio Science & Technology Co. Ltd), 400mM NaCl (AR/ACS Finar mw 58.44), 0.001M DTT (Merck) a ratio of 1:100 as illustrated in Table 2.

**Table 2.** Showing serial dilutions of recombinant viral HPV E6 protein using a protein diluent.

| **Tube Number** | **Concentration of E6 protein (mg/ml)** | **Concentration of E6 protein (pg/ml)** |
| --- | --- | --- |
| 1. | 0.5 | 500,000,000 |
| 2. | 0.005 | 5,000,000 |
| 3. | 0.00005 | 50,000 |
| 4. | 0.0000005 | 500 |
| 5. | 0.000000005 | 5 |
| 6. | 0.00000000005 | 0.05 |
| 7. | 0.00000000000005 | 0.0005 |
| 8. | 0.0000000000000005 | 0.000005 |
| 9. | 0.000000000000000005 | 0.00000005 |
| 10. | 0.00000000000000000005 | 0.0000000005 |

### Verification of signal amplification

The dot blot molecular technique was carried out using the serially diluted recombinant viral HPV E6 using unconjugated HPV 16 E6/18 E6-HRP (CP15) antibody and HPV 16 E6/18 E6- HRP(CP15)-AuNPs bioconjugated antibody.

### Determination of diagnostic accuracy of the cervical cancer testing nanokit using histopathology as the gold standard test

#### Protein extraction using RIPA Lysis method

Deidentified formalin fixed paraffin embedded archived biopsy specimens with confirmed histopathological findings were homogenized as follows. 10 μm of FFPE tissue sections were cut using microtomy with intervals of ethanol (Reagent grade ACS, ISO C_2_H_5_OH Scharlau) cleansing to avoid cross contamination and placed in 1.5 ml Eppendorf tubes. They were deparaffinized at room temperature in 500μl Xylene (Narcolab) for 10 minutes. The tissues were pelleted at 14000 × g for 3 minutes at 4°C, and deparaffinization repeated twice each time discarding the supernatant. The tissue pellets were rehydrated in decreasing graded series of ethanol ranging from absolute 100%, 90% and 70%. The tissue pellets were resuspended in ice-cold RIPA lysis extraction buffer (Cat No. R0020 Beijing Solarbio Science & Technology Co. Ltd.) with PMSF cocktail of protease inhibitors. The amount used was according to the size of the tissue in mg; 1ml for 100mg (26).The tissues were vortexed on ice 5 times 30 seconds each, then incubated on ice for 15 minutes with pipette mixing. The extracts were centrifuged at 14000 × g for 20min at 4°C to pellet the unsolubilized material. The supernatant was transferred to fresh tubes and stored at -20^0^C for downstream processes.

#### Testing the samples using the nanokit

About 2 μl of the cervical biopsy protein extracts were spotted onto the NC and MN paper strips at the center of the grid slowly ensuring about 2–4 mm diameter using narrow-mouth FinTip pipette tips (Thomas Scientific). The paper strips were air dried and non-specific sites blocked by soaking in 5% BSA in TBS-T in a 10 cm Petri dish for 1 hour at room temperature. The paper strips were incubated with HPV 16 E6/18 E6-HRP (CP15)-AuNPs bioconjugate to allow formation of an immune complex at a concentration of 4 μg/ml for 30 min at room temperature. The paper strips were washed three times with TBS-T (3 × 5 min) and incubated with TMB reagent. Blue color developed in 3 minutes.

#### Semi-quantitation of test results

Using the semi quantitative color code, the concentrations of E6 protein was determined and parameters used to plot the ROC Curve using easyROC curve analysis (ver. 1.3.1) as Table 3

**Table 3.** Showing the test sample estimate concentration, and the clinical sensitivity and 1-specificity used in plotting the ROC Curve.

| **Biopsy Identity** | **E6 Protein Concentration pg/ml** | **Nanokit Diagnosis** | **1-positive (Sum)** | **0-Negative (Sum)** | **Clinical Sensitivity (TPR =TP/(TP+FN))** | **1-specificity (FPR =FP/(FP+TN))** |
| --- | --- | --- | --- | --- | --- | --- |
| 348 | 50000000 | 1 | 1 | 0 | 0.025 | 0 |
| 375 | 50000000 | 1 | 2 | 0 | 0.05 | 0 |
| 586 | 50000000 | 1 | 3 | 0 | 0.075 | 0 |
| 140 | 50000000 | 1 | 4 | 0 | 0.1 | 0 |
| 336 | 50000 | 1 | 5 | 0 | 0.125 | 0 |
| 155 | 500 | 1 | 6 | 0 | 0.15 | 0 |
| 134 | 500 | 1 | 7 | 0 | 0.175 | 0 |
| 206 | 500 | 1 | 8 | 0 | 0.2 | 0 |
| 368 | 5 | 1 | 9 | 0 | 0.225 | 0 |
| 271 | 5 | 1 | 10 | 0 | 0.25 | 0 |
| 330 | 5 | 1 | 11 | 0 | 0.275 | 0 |
| 445 | 5 | 1 | 12 | 0 | 0.3 | 0 |
| 418 | 5 | 1 | 13 | 0 | 0.325 | 0 |
| 146 | 5 | 1 | 14 | 0 | 0.35 | 0 |
| 341 | 5 | 1 | 15 | 0 | 0.375 | 0 |
| 579 | 5 | 1 | 16 | 0 | 0.4 | 0 |
| 212 | 5 | 1 | 17 | 0 | 0.425 | 0 |
| 587 | 5 | 1 | 18 | 0 | 0.45 | 0 |
| 148 | 5 | 1 | 19 | 0 | 0.475 | 0 |
| 340 | 5 | 1 | 20 | 0 | 0.5 | 0 |
| 270 | 5 | 1 | 21 | 0 | 0.525 | 0 |
| 582 | 5 | 1 | 22 | 0 | 0.55 | 0 |
| 145 | 5 | 1 | 23 | 0 | 0.575 | 0 |
| 272 | 5 | 1 | 24 | 0 | 0.6 | 0 |
| 202 | 5 | 1 | 25 | 0 | 0.625 | 0 |
| 207 | 5 | 1 | 26 | 0 | 0.65 | 0 |
| 141 | 0.05 | 1 | 27 | 0 | 0.675 | 0 |
| 551 | 0.05 | 1 | 28 | 0 | 0.7 | 0 |
| 300 | 0.05 | 1 | 29 | 0 | 0.725 | 0 |
| 391 | 0.05 | 1 | 30 | 0 | 0.75 | 0 |
| 338 | 0.05 | 1 | 31 | 0 | 0.775 | 0 |
| 147 | 0.0005 | 0 | 31 | 1 | 0.775 | 0.1 |
| 630 | 0.0005 | 1 | 32 | 1 | 0.8 | 0.1 |
| 430 | 0.0005 | 1 | 33 | 1 | 0.825 | 0.1 |
| 571 | 0.0005 | 1 | 34 | 1 | 0.85 | 0.1 |
| 139 | 0.0005 | 1 | 35 | 1 | 0.875 | 0.1 |

**Table 3. Continued**
| **Biopsy Identity** | **E6 Protein Concentration pg/ml** | **Nanokit Diagnoses** | **1-positive (Sum)** | **0-Negative (Sum)** | **Clinical Sensitivity (TPR = TP/(TP+FN))** | **1-specificity (FPR=FP/(FP+TN))** |
| --- | --- | --- | --- | --- | --- | --- |
| 585 | 0.0005 | 1 | 36 | 1 | 0.9 | 0.1 |
| 265 | 0.000005 | 0 | 36 | 2 | 0.9 | 0.2 |
| 184 | 0.000005 | 0 | 36 | 3 | 0.9 | 0.3 |
| 156 | 0.000005 | 0 | 36 | 4 | 0.9 | 0.4 |
| 187 | 0.000005 | 0 | 36 | 5 | 0.9 | 0.5 |
| 425 | 0.000005 | 0 | 36 | 6 | 0.9 | 0.6 |
| 263 | 0.000005 | 0 | 36 | 7 | 0.9 | 0.7 |
| 135 | 0.000005 | 0 | 36 | 8 | 0.9 | 0.8 |
| 386 | 0.000005 | 0 | 36 | 9 | 0.9 | 0.9 |
| 355 | 0.000005 | 0 | 36 | 10 | 0.9 | 1 |
| 577 | 0.000005 | 1 | 37 | 10 | 0.925 | 1 |
| 549 | 0.000005 | 1 | 38 | 10 | 0.95 | 1 |
| 576 | 0.000005 | 1 | 39 | 10 | 0.975 | 1 |
| 583 | 0.000005 | 1 | 40 | 10 | 1 | 1 |
|  | Subjects with Disease |  | 40 |  |  |  |
|  | Subjects without Disease |  | 10 |  |  |  |

Nanokit diagnosis
1 Indicates positive
0 Indicates negative
TPR True positive rate
FPR False positive rate

## Results

### Evaluation of immune sensing mechanism of the HPV 16 E6/18 E6-HRP (CP15) -AuNPS bioconjugate

The antibody bioconjugate formed proved its bio-functionality on HPV E6 recombinant protein using dot blot molecular technique as observed by colour change from colourless to blue. Differing and serial dilutions of recombinant HPV E6 protein were obtained for the dot blot assay from an initial concentration of 0.5mg/ml of protein.

### Immune sensing mechanism

The principle of antibody antigen reaction was investigated successfully, as the gold conjugated antibody possessed the ability to recognize and bind specific E6 oncoprotein macromolecule as illustrated on Plate 1.

**Plate 1.**
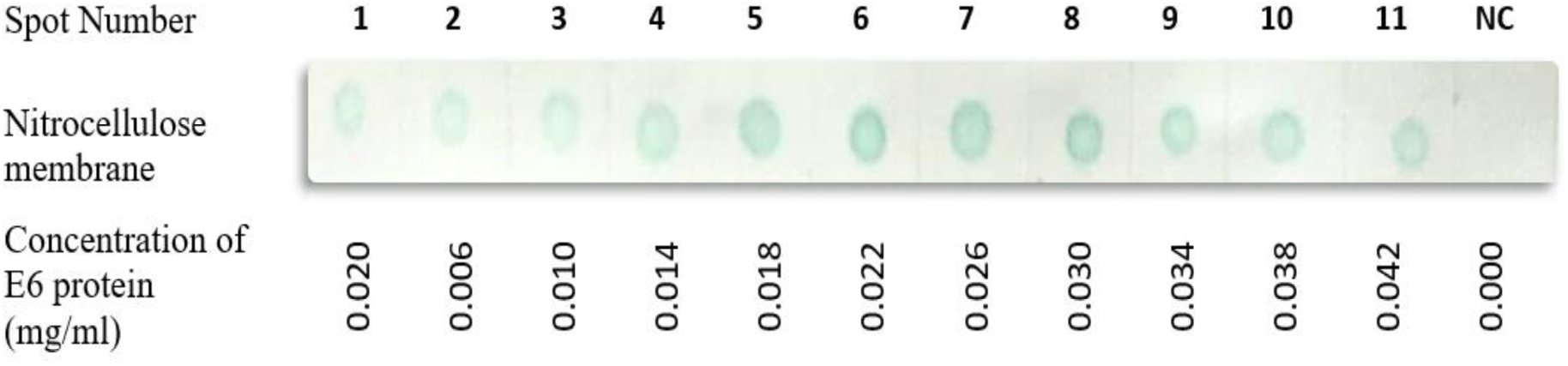
Showing the Anti HPV 16/18 E6-HRP-AuNPs Conjugate Dot-Blot reaction with recombinant viral HPV E6 purified protein. A colour change to blue (positive result) was generated by catalytic oxidation of peroxidase enzyme on 3,3’,5,5’-Tetramethylbenzidine (TMB) substrate. It shows an increase in signal from spot 1 to 8 due to increasing concentration of E6 protein. 9–11 varied due to possible bioconjugate instabilities. NC is the negative control containing protein diluent buffer.

### Determination of Limit of Detection

The lowest detectable E6 oncoprotein amount was obtained at 0.0005 pg/ml indicating a highly sensitive test, see Plate 2.

**Plate 2.**
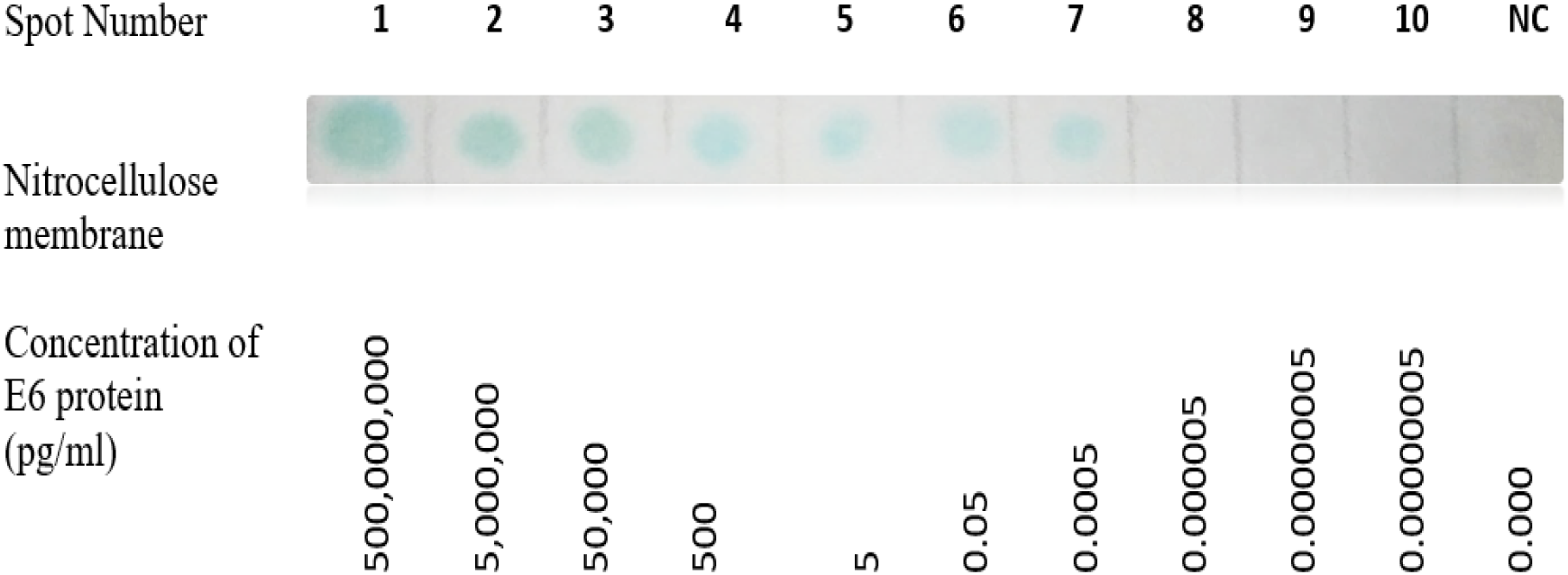
Showing the Anti HPV 16/18 E6-HRP-AuNPS Conjugate Dot-Blot reaction with serially diluted recombinant viral HPV E6 purified protein. A colour change to blue was generated by catalytic oxidation of peroxidase enzyme on 3,3’,5,5’-Tetramethylbenzidine (TMB) substrate. The lowest concentration obtained is 0.0005pg/ml (Spot 7).

### Verification of signal amplification

A comparison of immune sensing mechanisms between unconjugated antibody and conjugated antibody with the recombinant viral HPV E6 purified protein showed E6 oncoprotein detection is improved by conjugating the antibody to gold nanoparticles. It shows an increased signal for gold conjugated antibody compared to unconjugated antibody. Spot 1–7 show a decrease in signal due to decreasing concentration of E6 protein after serial dilution of the HPV E6 protein as shown on Plate 3.

**Plate 1:**
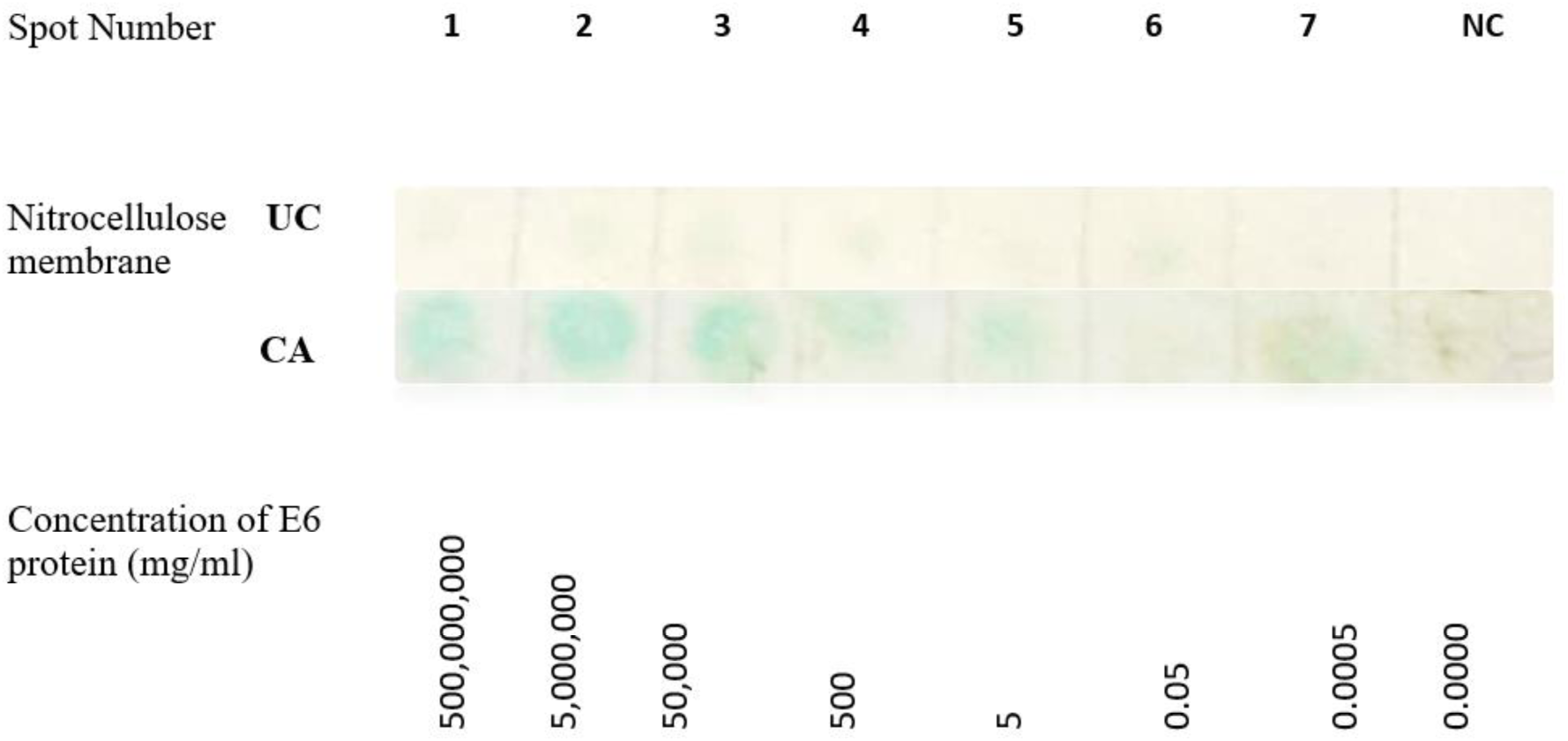
Showing the Unconjugated Antibody (UA) and Conjugated Antibody (CA) Dot-Blot reactions with differing concentrations of recombinant viral HPV E6 purified protein. A colour change to blue (positive result) was generated by catalytic oxidation of peroxidase enzyme on 3,3’,5,5’-Tetramethylbenzidme (TMB) substrate. NC is the negative control containing protein diluent buffer.

### Determination of diagnostic accuracy of the cervical cancer testing nanokit using histopathology as the gold standard test

Accuracy and validity of a diagnostic test was measured in reference to a gold standard test for diagnosis of cervical cancer the histopathology test. The archived biopsy samples had confirmed pathology in grades of cervical cancer progression. This included; the normal tissue, mild dysplasia (CIN1 & CIN2) tissue, CIN3 tissue, CIN4 tissue, microinvasive and macroinvasive cervical samples. E6 protein increases with progression from mild dysplasia to macroinvasive cancer and hence, the blue colour development of the nanokit increases from mild dysplasia to macroinvasive cervical cancer, see Plate 4.

**Plate 2:**
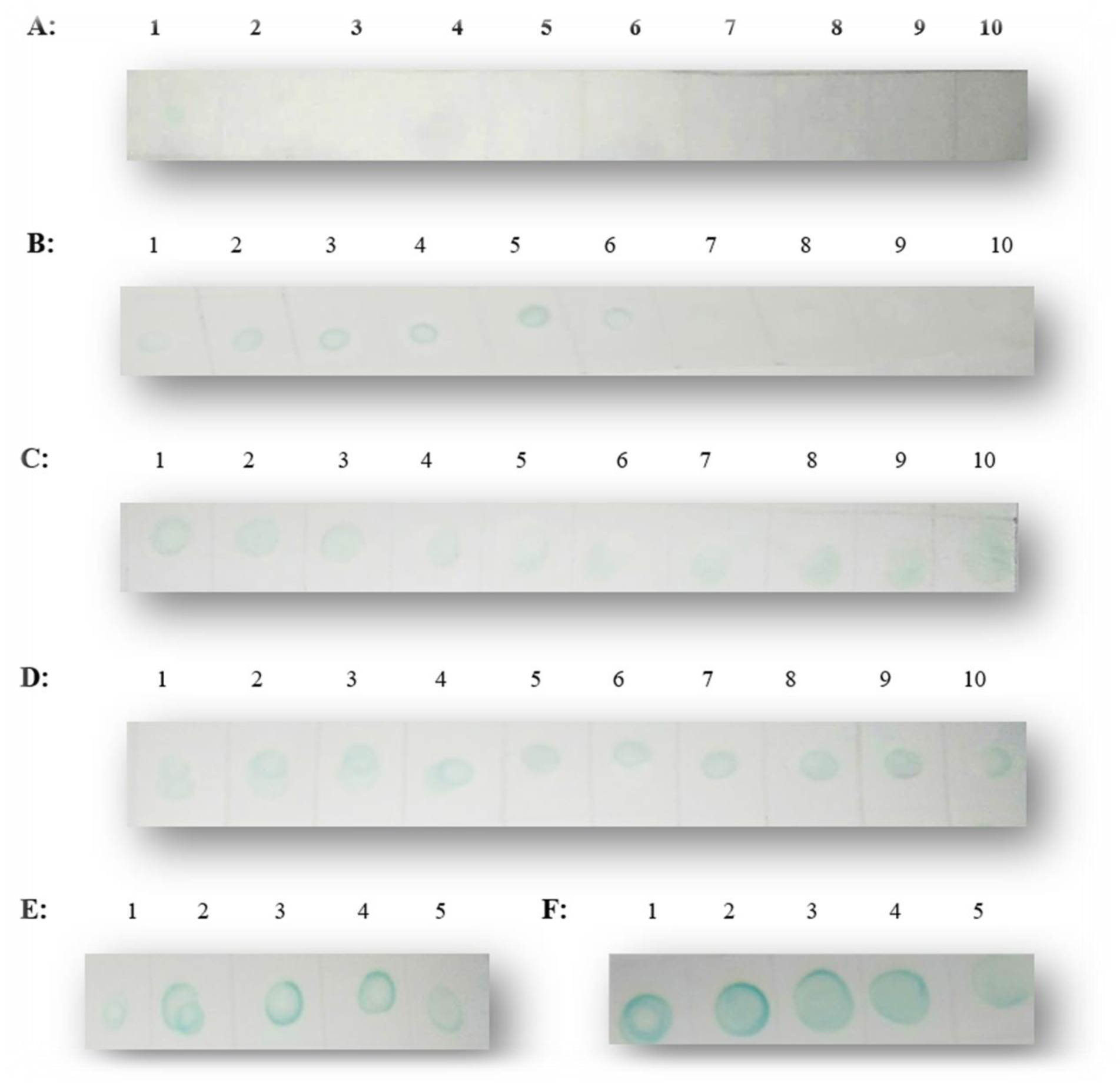
Showing HPV 16 E6/18 E6-HRP (CP15)-AuNPS Conjugate Dot-Blot reactions with Formalin Fixed Paraffin Embedded, FFPE tissue lysates of confirmed pathology. **(A)** The normal cervical biopsy negative reaction, spot 1 however had a weak positive reaction (False positive); **(B)** Mild Dysplasia (CIN1 & CIN 2) indicated weak positive reaction with 4 false negatives spot 7–10; **(C)** CIN3 positive reaction; **(D)** CIN4 positive reaction; **(E)** Microinvasive positive reaction and **(F)** Macroinvasive positive reaction (This recorded the strongest signal). The colour change to blue (positive result) was generated by catalytic oxidation of peroxidase enzyme on 3,3’,5,5’-Tetramethylbenzidine substrate.

### The 2×2 contingency table

This is a table showing the distribution of variables in rows and others in columns, used to study the correlation between these variables. See Table 4.

**Table 4:** 2×2 table for calculation of sensitivity and specificity with groups of subjects divided with reference to a gold standard test (column) and categories according to test (rows). **TP** refers to True Positive, **FP** refers to False Positive, **FN** refers to False Negative while **TN** refers to True negative.

|  | Subjects with the disease | Subjects without the disease | Totals |
| --- | --- | --- | --- |
| Positive | TP (36) | FP (1) | 37 |
| Negative | FN (4) | TN (9) | 13 |
| Totals | 40 | 10 | 50 |

**Sensitivity** is the probability of getting a positive test result in subjects with the disease (T+|B+).

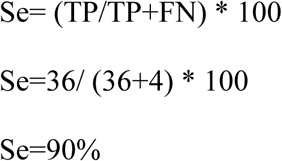

**Specificity** is the probability of a negative test result in a subject without the disease (T-|B-)

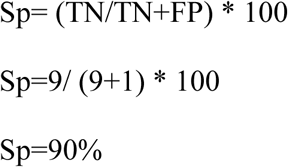

**Likelihood Ratio** is the ratio of expected test result in subjects with a certain disease to the subjects without the disease.

Likelihood Ratio for positive test results, LR+= sensitivity / (1-specificity)

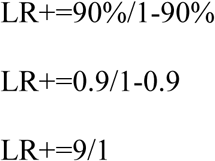

Likelihood Ratio for negative test results, LR- = (1-sensitivity) / specificity

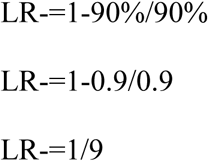

**Positive predictive value (PPV)** defines the probability of having the disease of interest in a subject with positive result (B+|T+)

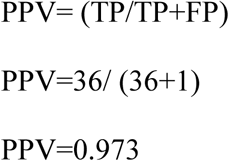

**Negative predictive value (NPV)** describes the probability of not having a disease in a subject with a negative test result (B-|T-).

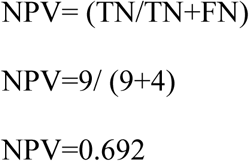

### The Receiver Operating Characteristic (ROC) Curve

A receiver operating characteristic (ROC) curve, being a plot of test sensitivity as the y coordinate versus its 1-specificity or false positive rate (FPR) as the x coordinate was used to evaluate the quality of the developed nanokit (27). The coordinates used are illustrated on Table 5.

**Table 5:** Showing the ROC Coordinates used to derive the ROC curve.

| **E6 oncoprotein Marker** | **Cutpoint** | **FPR(1-specificity)** | **TPR (Sensitivity)** |
| --- | --- | --- | --- |
| **Concentration** | **-Inf** | **0** | **0** |
| **Concentration** | **5e-06** | **0.1** | **0.9** |
| **Concentration** | **5e-04** | **0.225** | **1** |
| **Concentration** | **0.05** | **0.35** | **1** |
| **Concentration** | **5** | **0.8** | **1** |
| **Concentration** | **500** | **0.875** | **1** |
| **Concentration** | **50000** | **0.9** | **1** |
| **Concentration** | **5e+07** | **1** | **1** |
| **Concentration** | **Inf** | **1** | **1** |

**Fig 1.**
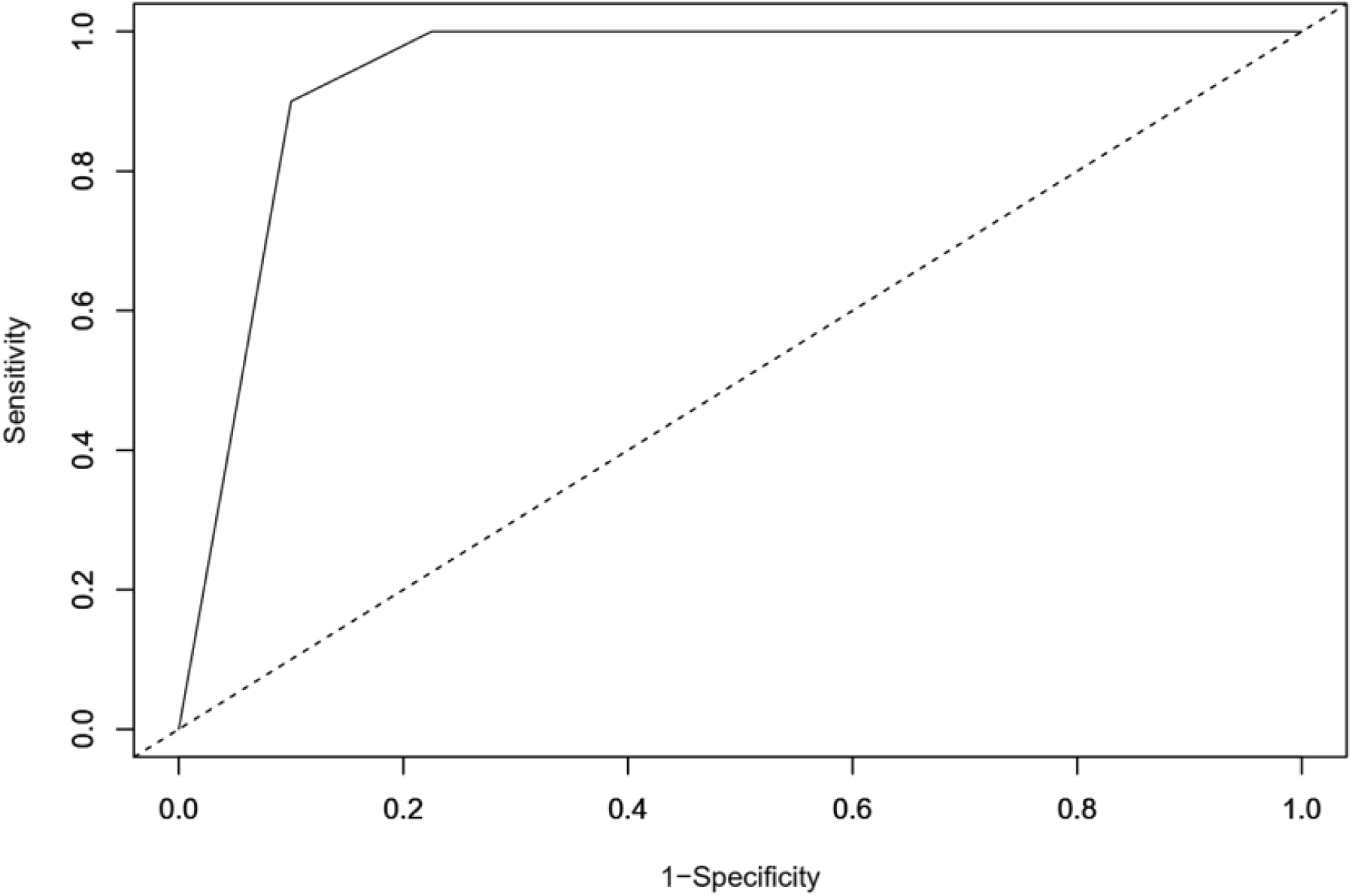
Showing the ROC curve for the E6 oncoprotein nanokit test on cervical cancer biopsy formalin Fixed paraffin embedded tissues.

### Principle of the nanokit developed in this study

The nanokit uses the direct ELISA-based immunoassay principle as illustrated in Figure 2.

**Figure 2.**
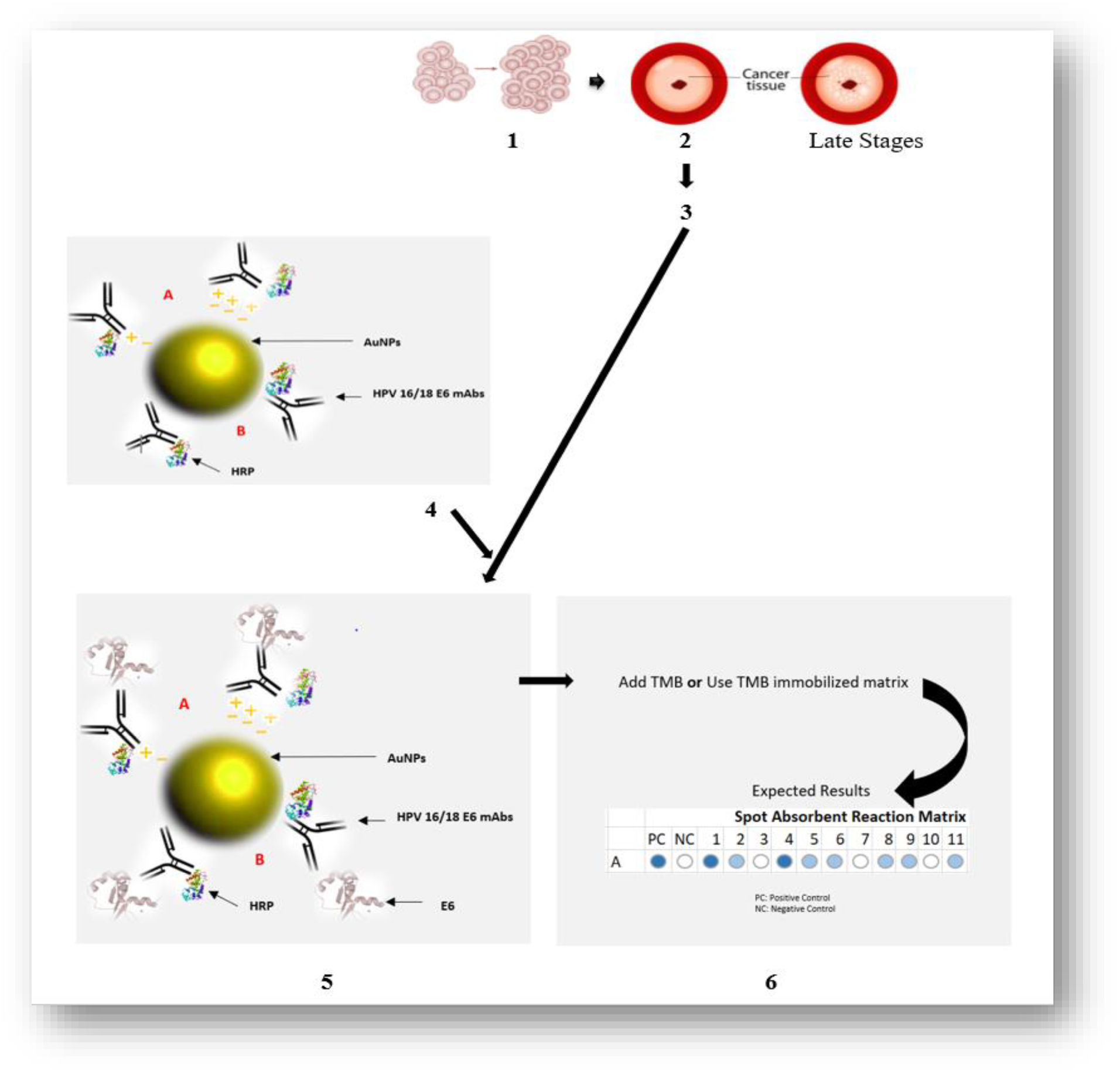
Illustrating the paper-based nanokit principle. (1) Malignant transformation of cells due to HPV infection, (2) Dysplasia becomes apparent at the cervical tissue during early stage (3) During this early stage Early oncogenic protein, E6 is released (4) the HPV 16/18 E6-HRP (CP 15)-AuNPS bioconjugate that was developed in the study is reacted with E6 oncogenic protein yielding (5) an immune complex (6) Addition of TMB or deposit on TMB immobilized template yields the colorimetric results

## Discussion

### Evaluation of immune sensing mechanism of the HPV 16 E6/18 E6-HRP (CP15) -AuNPS bioconjugate

HPV 16 E6/18 E6-HRP (CP15) -AuNPS bioconjugate was evaluated for specific binding to E6 oncoprotein analyte using the dot blot molecular technique (25). A dot blot is a technique similar to western blot as they both utilize antibodies to identify specific membrane bound proteins. It however, doesn’t require gel electrophoretic protein separation hence cannot determine the molecular weight of a protein or to discriminate between different protein. This study utilized dot blots to determine whether the gold conjugated antibody detection system will work effectively to identify the protein in the tissue lysate crude preparations used. The study also used HRP directly labeled primary antibody to maximize on specific binding, reduce reaction time and enhance data quality through assay simplification (28).

The bioconjugate possesses immune sensing mechanism for the E6 protein yielding blue colorimetric results generated by catalytic oxidation of hydrogen peroxidase enzyme on 3,3’,5,5’- Tetramethylbenzidine (TMB) substrate.

In this study, the gold nanoparticles conjugated antibody exhibited an amplified immunological signal with the recombinant viral HPV E6 protein than the unconjugated antibody. This is because gold nanoparticles possess optical properties at nanoscale measurements that also enhance the colorimetric signal produced by the peroxidase activity of horseradish peroxidase (HRP) enzyme with TMB.

The dot blot molecular technique carried out with serially diluted recombinant viral HPV E6 protein yielded a semiquantitative color code with different grades of colorimetric signal. This is used to partially estimate the amount of E6 oncoprotein expressed in test samples. This nanokit assay obtained the lowest amount of analyte (Limit of Detection (LOD)) as 0.0005pg/ml.

### Determination of diagnostic accuracy of the cervical cancer testing nanokit using histopathology as the gold standard test (reference standard)

This study provides a significantly high and reliable diagnostic accuracy for precancerous lesions mild dysplasia, (CIN3), (CIN4), microinvasive and macroinvasive cancerous lesions using 2×2 contingency table analysis of parameters of diagnostic test quality. The nanokit obtained values that fall within the upper and lower limits for a quality diagnostic test are outlined in the Table 5.1 (23,29).

**Table 6:** Showing that the parameters of diagnostic test quality for the nanokit fall within the recommended lower and upper limits.

|  | **Nanokit Value** | **Lower Limit** | **Upper Limit** |
| --- | --- | --- | --- |
| **Sensitivity** | 0.900 | 0.555 | 0.997 |
| **Specificity** | 0.900 | 0.763 | 0.972 |
| **Positive Predictive Value** | 0.692 | 0.446 | 0.990 |
| **Negative Predictive Value** | 0.973 | 0.833 | 0.993 |
| **Positive Likelihood Ratio** | 9.000 | 3.472 | 23.327 |
| **Negative Likelihood Ratio** | 0.111 | 0.017 | 0.715 |

A receiver operating characteristics (ROC) curve displays the trade-offs between sensitivity and specificity for a test on a continuous or ordinal scale. The ROC curve plotted shows a consistent plot nearer to the left-hand side and the top, which is the most preferred (29). The area under the ROC curve (AUROC) of the nanokit test was 0.93875 which is within the lower limit of 0.88283 and upper limit of 0.99467 of a good diagnostic test (23). AUROC is a measure of the overall performance of a diagnostic test and is interpreted as the average value of sensitivity for all possible values of specificity. It can take any value between 0 and 1, since both the x and y axes have values ranging from 0 to 1. The closer AUROC is to 1, the better the overall diagnostic performance of the test, as an ideal test has AUROC value of 1 (27,31).

World Health Organization (WHO) Sexually Transmitted Diseases Diagnostics Initiative (SDI) defines the diagnostic test quality using the ASSURED criteria. This is an acronym for Affordable, Sensitive, Specific, User-friendly, Rapid and robust, Equipment-free and Deliverable to end-users (32). The paper-based nanokit developed in this proof of concept study is sensitive, specific and rapid. With more optimization and verification, the nanokit presents a promising future in achieving the ASSURED criteria.

One of the study limitations was insufficiency of one FFPE biopsy sample after deparaffinization, thus the determination of diagnostic accuracy utilised (n=50) samples instead of the collected number of samples (n=51). Additionally, some test reactions were fairy reproducible due to possible bioconjugate instabilities.

## Conclusion

The study showed that the HPV 16 E6/18 E6-HRP(CP15) - AuNPs conjugate possesses immune sensing mechanism for HPV 16/18 Early 6 (E6) oncoprotein produced during early stages of cervical cancer progression and that the conjugation enhances the detection of E6 oncoprotein. The study also showed a commendable diagnostic accuracy of the nanokit using histopathology as the gold standard.

## Recommendations

The following points are recommended for further work so as to maximize the benefits from this study:

I. The HPV 16 E6/18 E6-HRP (CP15)-AuNPs conjugate be used in further testing on freshly collected cervico-vaginal specimens and more optimization to enable its nanofabrication.
II. Since the bioconjugate exhibited instabilities in some reactions, covalent bioconjugation be considered in subsequent productions.
III. To verify absence of unspecific binding (cross-reactions) of gold nanoparticle antibody bioconjugate to other similar macromolecules resulting from co-infections of HPV with other Viral species or normal flora, a closely related viral particle should be utilised to serve as control.
IV. A negative control protein known not to react with the antibody thereof should also be utilized.

## Data Availability

All data as pertains to the manuscript is available through all cited references.

## Acknowledgements

We would like to express our gratitude to the staff at Molecular Biology and biotechnology PAUSTI Laboratory and University of Nairobi Histopathology laboratory for their support and creating a conducive environment in which the lab investigations were carried out.

